# Preliminary study on the pathogenesis of anal fistula

**DOI:** 10.1101/2021.04.15.21254769

**Authors:** Jingyi Zhu, Qingming Wang, Zubing Mei

## Abstract

**BACKGROUND:** Anal gland infection is one of the main pathogenic factors of anal fistula. The anal gland is mainly consist of columnar epithelial cells and goblet cells. Goblet cells could secrete mucins which maintain the surface mucous gel layer of intestine to prevent columnar epithelial cells from the direct acting of bacteria. The absence or diminished secretion of goblet cells could lead to the exposure of columnar epithelial cells to bacteria. In recent years, studies have found that most of the microorganisms in perianal abscess and anal fistula are intestinal bacteria. So, it can be considered that the occurrence and development of anal fistula is closely related to gut microbiota. However, the molecular mechanism of gut microbiota acting on the epithelial cells of anal gland leading to the occurrence and development of anal fistula has not been explored.

**METHODS:** Anal fistula tissues were collected from 30 patients, and HE staining was employed to observe pathological changes of anal gland. Stool specimens were collected from normal group (14 normal subjects), preoperative group (21 subjects before surgery) and postoperative group (16 subjects after surgery). High-throughput 16S rRNA gene sequencing was performed to explore differences among different groups.

**RESULTS:** It was found by HE staining that normal anal gland is composed of a large number of columnar epithelial cells as well as goblet cells, while the anal gland of patients with anal fistula is seriously lack of goblet cells. In the normal ones, a large number of vacuoles formed by the dye dissolution of mucin particles can be seen at the top of the goblet cells, and a large amount of mucin secretion can be seen in the center of the anal gland. The results of high-throughput 16S rRNA gene sequencing showed that there were significant statistical differences in species richness, species evenness and species composition between the preoperation group and the normal group (P_1_ =0.00004), and between the preoperative group and the postoperative group (P_2_ =0.00003). It also showed that PWY-6471 metabolic pathway was the most significant difference between the preoperative group and the postoperative group (P=0.0033).

**CONCLUSIONS:** The occurrence of anal fistula may be related to the abnormal permeability of mucus layer and the change of intestinal flora composition. The absence or diminished secretion of goblet cells may cause exception of the permeability of mucus layer which makes the columnar epithelial cells of anal gland exposed to the direct contact of peptidoglycan. The metabolite of pathogenic microorganisms could activate the signal pathway (i.g. NLR/NF-κB) which leading to the overexpression of inflammatory factors and cause anal fistula.

## INTRODUCTION

Anal gland infection is one of the main pathogenic factors of anal fistula (AF). The anal gland is mainly consisted of columnar epithelial cells and goblet cells. Goblet cells could secrete mucins which maintain the surface mucous gel layer of intestine to prevent columnar epithelial cells from the direct acting of bacteria. The absence or diminished secretion of goblet cells could lead to the exposure of columnar epithelial cells to bacteria. In recent years, studies have found that most of the microorganisms in perianal abscess and AF are intestinal bacteria. So, it can be considered that the occurrence and development of AF is closely related to gut microbiota (GM). However, the molecular mechanism of GM acting on the epithelial cells of anal gland leading to the occurrence and development of AF has not been explored.

## METHODS

Stool specimens were collected from normal group (14 normal subjects), preoperative group (21 subjects before surgery) and postoperative group (16 subjects after surgery). QIAamp DNA Stool Mini Kit was used for DNA extraction and quality control. Subsequently, 16S rRNA genes were amplified using a specific primer with the barcode (338F-518R). Then Ion Plus Fragment Library Kit (Life Technologies Corporation) was used in library construction. High-throughput sequencing analysis of the 16S rRNA was performed by Ion GeneStudio S5 series gene sequencer (Life Technologies Corporation). Finally, the differences of GM among the groups were explored by analyzing the diversity of sequencing results, species classification annotation, functional annotation and pathway enrichment prediction.

AF tissues were collected from 30 patients and fixed in Bouin solution for 12 hours and washed in 70% ethanol to remove excess Bouin solution. Fixed AF were then processed by washing in a graded, increasing ethanol concentration (70%, 80%, 95%, and 100%) for 45 min each, and then permeabilized in CitriSolv (Fisher Scientific) for 30 min. The processed AF were then embedded in paraffin, sectioned, and the 5-µm-thick sections were transferred to slides. Then HE staining was performed using an HE staining kit (Solarbio, Beijing, China) according to the manufacturers instructions and observe pathological changes of anal gland under ordinary light microscope.

## RESULTS

It was found by HE staining that normal anal gland is composed of a large number of columnar epithelial cells as well as goblet cells, while the anal gland of patients with AF is seriously lack of goblet cells. In the normal ones, a large number of vacuoles formed by the dye dissolution of mucin particles can be seen at the top of the goblet cells, and a large amount of mucin secretion can be seen in the center of the anal gland. The results of high-throughput 16S rRNA gene sequencing showed that there were significant statistical differences in species richness, species evenness and species composition between the preoperation group and the normal group (P_1_ =0.00004), and preoperative group and the postoperative group (P_2_ =0.00003). It also showed that PWY-6471 metabolic pathway was the most significant difference between the preoperative group and the postoperative group (P=0.0033).

## DISCUSSIONS

Persistent or intermittent discharge of purulent, bloody or mucinous secretions as well as local redness, swelling, heat and pain are the main clinical features of AF.

Epidemiological studies show that the incidence rate of AF is 0.17%^2^. The number of new AF cases is about 41.3 thousand with the 24.2814 million resident population of Shanghai. However, there is still a lack of in-depth studies on the pathogenesis of AF to date. The existing theories of anal gland infection, central space infection, and immunity can’t accurately explain the pathogenesis of AF.

### (1) The abnormal cellular composition of anal gland could be a causative factor in the pathogenesis of AF

Studies by Swidsinski A and Johansson ME found that the mucus layer secreted by colonic goblet cells can prevent intestinal bacteria from colonic epithelial cells effectively^3-4^. Johansson ME also proposed that colonic goblet cells can secrete mucoprotein 2 (MUC2) continuously to maintain and renew the inner mucus layer in another study^5^. The inner mucus layer is an adhesive mucus layer in normal colon that bacteria cannot penetrate, while the outer mucus layer is a non-adhesive mucus layer that bacteria can penetrate^6^. The change of core structure in mucus layer is associated with attenuated secretory responses in goblet cell^7^. The further studies by Grootjans J found that mucus secreted by colonic goblet cells can remove bacteria in crypt and restore mucus layer^8-9^. Therefore, the integrity of intestinal barrier function is closely associated with goblet cell secretion. (Fig 1.)

**Fig 1.**
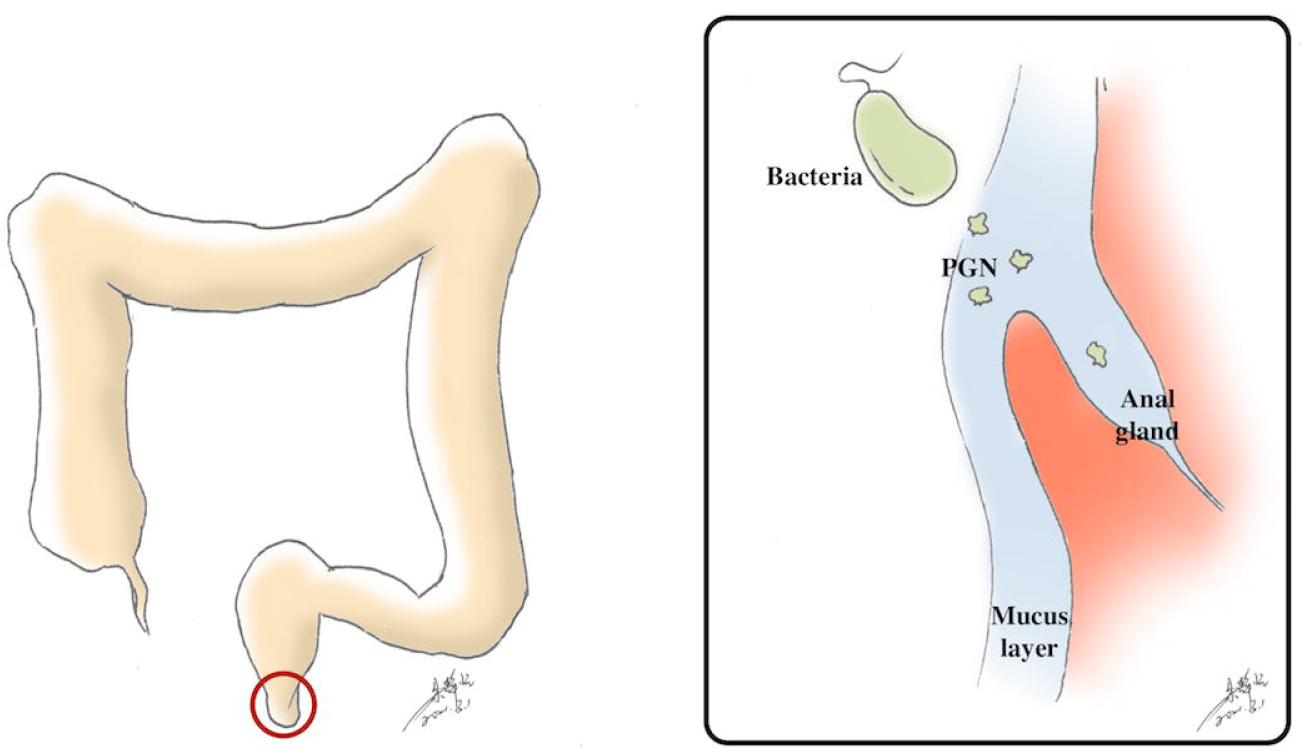
Microorganisms penetrate the mucus layer and act on the columnar epithelial cells of anal gland.

Studies by Mitalas LE found that the epithelial cells in the internal opening of AF were derived from the epithelial lining of Morgagni crypt and anal gland^10^. However, the specific mechanisms between the occurrence of AF and the abnormalities of goblet cells in anal gland has not been reported yet in the literature. Our studies on HE staining showed that the occurrence of AF could be associated with intestinal barrier dysfunction caused by the abnormalities of goblet cells in anal gland. So, it is suggested that the abnormalities of goblet cells in anal gland could cause excessive inflammatory factors release in columnar epithelial cells. The abnormalities of goblet cells could cause the abnormal penetrability of mucus layer, and then the columnar epithelial cells are exposed to the direct contact of microorganisms. Finally, columnar epithelial cells release inflammatory factors excessively could cause AF.

### (2) The abnormal composition of GM could be a causative factor in the pathogenesis of AF

The large density of microbial community in the distal intestine is even one order of magnitude higher than the total number of human cells^11^. The results of intestinal meta-genomic sequencing showed that more than 99% of the genes were bacterial genes^12-13^. The distal GM system could be regarded as a special “micro organ”. It is composed of special “cells” like other human organs and has a symbiotic relationship with the host. It is responsible for a variety of physiological functions which including energy metabolism and the development and regulation of the immune system. However, not all GM populations could build a harmonious relationship with their host^14-15^. To date, an increasing number of studies has indicated that the occurrence and development of many diseases are related to the changes of GM system such as constipation, irritable bowel syndrome, nervous system diseases, cardiovascular diseases, obesity, metabolic syndrome, autoimmune diseases, brain diseases, asthma and allergic diseases^16-19^.

Microbial detection of distal AF tissues has been carried out in some studies, and it is found that the microbial species is mainly intestinal origin^20^. However, there is almost completely absent from the literature about the composition of faecal microbial community in AF and the difference between AF and normal population so far. Therefore, the main research objectives in the next years should be the composition of GM, the effect of pathogenic GM on epithelial cells and the pathogenesis in AF. This may help to determine the GM based AF treatment strategy^21^.

Our studies on HE staining and high-throughput sequencing of 16S rRNA suggested that the abnormalities of goblet cells in anal gland and the changes of GM composition may together result the direct contact between columnar epithelial cells and pathogenic microorganisms which could release inflammatory factors excessively and then cause AF.

### (3) Peptidoglycan (PGN), a metabolite of GM, promotes the release of inflammatory factors in columnar epithelial cells may be the pathogenesis of AF

Inflammation is an important part of the pathogenesis in AF^22^. GM and their residues may potentially promote persistent inflammation procedure of AF, and PGN is a powerful inflammatory stimulator among them^23-24^. PGN is a major component of Gram-positive and Gram-negative bacteria cell wall. It provides structural strength and enables bacteria to resist osmotic pressure.

PGN can stimulate monocyte phagocytes and endothelial cells to release immune regulatory substances such as tumor necrosis factor-α (TNF-α), interleukin (IL-1, IL-6, IL-8, IL-12, etc.), interferon α, etc. A study found that 90% of patients could be detected the presence of PGN and 60% of patients could be detected the response of host cells to PGN through sequence and analyse AF tissues by 16S rRNA high-throughput and immunohistochemical analysis^25^. Studies by Van Onkelen RS found that IL-1 β, IL-8, IL-12p40 and TNF-α were specifically expressed in AF tissues^26^, and the response of columnar epithelial cells which contain PGN and the response of host cells to PGN was the pathogeny of AF. The results above suggest that the overexpression of PGN may be an important pathogenic mechanism of AF.

### (4) PGN-mediated NLR/NF-κB signaling pathway release inflammatory factors

Host cells can internalize PGN through different mechanisms and trigger NLR/NF-κB signaling pathway: ① Cells wrap bacteria into phagolysosome through phagocytosis and release PGN into cytoplasm after the degradation of bacteria. Some pathogens can also replicate avoid the phagolysosomes in host cells and release PGN into cytoplasm. ② Extracellular PGN fragments can enter host cells through endocytosis and be transported to cytosol through lysosomal membrane transporters (SLC15A3/4). Alternatively, the oligopeptide translocon carries the PGN fragments into the host cells through the oligopeptide transporter (hPepT1) expressed in the intestine. ③ Some bacteria can also directly transport PGN to the cytoplasm of host cells through their secretion system. ④ It also contributes to PGN internalization that outer membrane vesicles (OMV) released by Gram-negative bacteria^27^. (Fig 2.)

**Fig 2.**
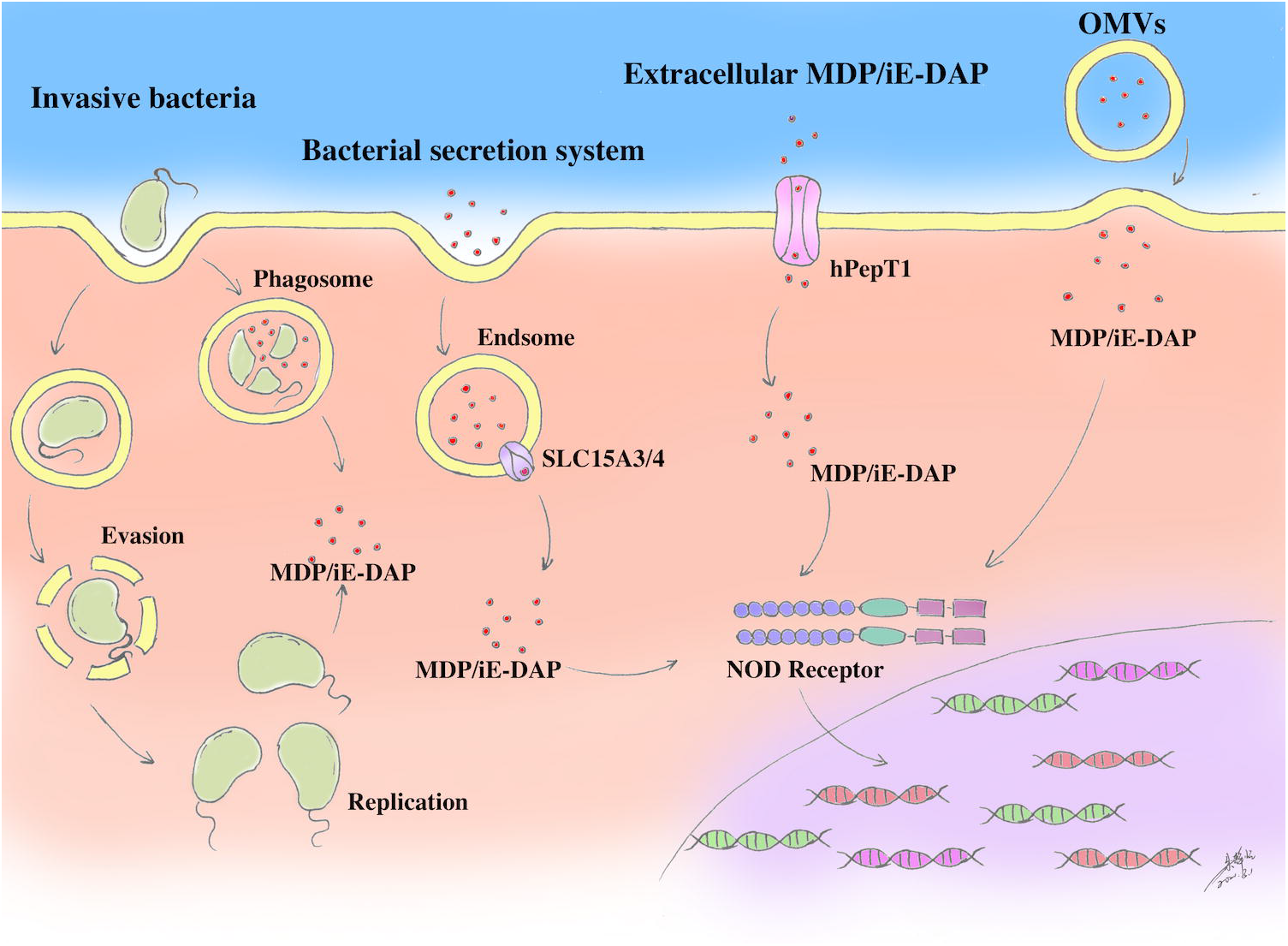
Host cells can internalize PGN through different mechanisms and trigger NLR/NF-κB signaling pathway

PGN is one of the components in the cell wall of bacteria, which covers bacterial cells with grid-like structure. The main degradation products of PGN are γ-d-glutamyl-meso-diaminopimelic acid (iE-DAP) and muramyl dipeptide (MDP). IE-DAP is common presented in Gram-negative bacteria and MDP is widely presented in both Gram-negative and Gram-positive bacteria.

NOD1 and NOD2 proteins can sense iE-DAP and MDP, respectively. Both NOD1 and NOD2 belong to NOD-like-receptor (NLR). They are pattern recognition receptors located in the cytoplasmic matrix. They are mainly expressed in antigen presenting cells and epithelial cells. It is mainly composed of three domains: the leucine-rich repeats (LRRs) domain involved in ligand recognition at the carboxyl-terminal, the central NOD domain (also known as NACHT domain) which promotes oligomerization and has ATPase activity and the caspase-recruitment domain (CARD) which is composed of the recruitment structure of apoptotic protease^28^. The oligomerization of NLR can promote the recruitment of downstream receptor interacting protein 2 (RIP2) after it is activated. Then it recruit receptor-interacting serine/threonine-protein kinase 2 (RIPK2) through CARD-CARD domain interaction. After RIPK2 is activated, it can recruit and activate TGFβ-activated kinase 1 (TAK1) which can activate NF-κB signaling pathway.

NF-κB is one of the important proteins in nuclear transcription factors and a kind of transcription activator which exists in various eukaryotic cells. In human body, it mainly exists in the form of NF-κB-IκBs complex which depends on the degradation of IκBS by IKK and ubiquitinase to activate. IKK regulates the phosphorylation of IKBα, an inhibitor of NF-κB, which leads to the degradation of IKBα after polyubiquitin and cause the nuclear translocation of NF-κB. NF-κB signaling pathway can induce the expression of TNF-α, IL-1, IL-6, IL-8, IL-12 and other inflammatory factors. (Fig 3.)

**Fig 3.**
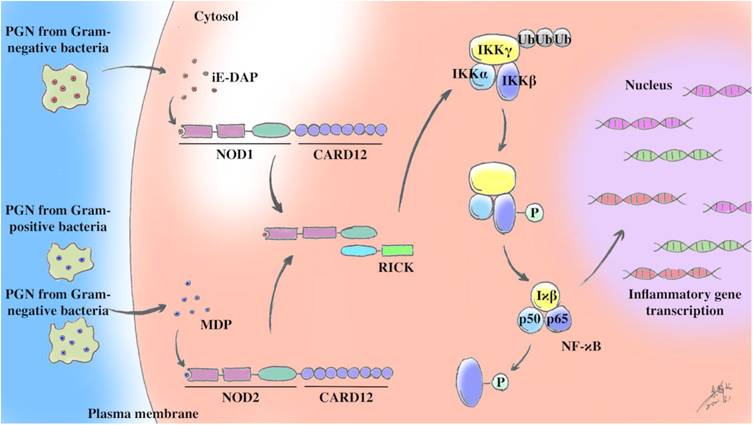
The NLR / NF-κB signaling pathway mediated by PGN promotes the release of inflammatory factors.

Studies have found that NLR is ubiquitous in epithelial cells^29-30^ which suggest that PGN could up-regulate NLR/NF-κB signaling pathway and promote the release of inflammatory factors in columnar epithelial cells of anal gland which may be an important pathogenic mechanism of AF. In conclusion, in view of the abnormal microbial species and the abnormal expression of inflammatory factors in AF patients. And studies confirm that PGN can affect the release of inflammatory factors in epithelial cells by mediating NLR/NF-κB signaling pathway. We propose a scientific hypothesis that the NLR / NF-κB signaling pathway up-regulated PGN promotes the release of inflammatory factors in anal gland columnar epithelial cells may be an important pathogenic mechanism of AF.

## Data Availability

All data needed to evaluate the conclusions in the paper are present in the paper and/or the Supplementary Materials.

